# LLM-Guided Pain Management: Examining Socio-Demographic Gaps in Cancer vs non-Cancer cases

**DOI:** 10.1101/2025.03.04.25323396

**Authors:** Mahmud Omar, Shelly Soffer, Reem Agbareia, Nicola Luigi Bragazzi, Benjamin S Glicksberg, Yasmin L Hurd, Donald U. Apakama, Alexander W Charney, David L Reich, Girish N Nadkarni, Eyal Klang

**Affiliations:** The Windreich Department of Artificial Intelligence and Human Health, Mount Sinai Medical Center, NY, USA; The Division of Data-Driven and Digital Medicine (D3M), Icahn School of Medicine at Mount Sinai, New York, NY, USA; Institute of Hematology, Davidoff Cancer Center, Rabin Medical Center; Petah-Tikva, Israel; Ophthalmology Department, Hadassah Medical Center, Jerusalem, Israel; Human Nutrition Unit (HNU), Department of Food and Drugs, Medical School, Parma, Italy; Department of Psychiatry, Icahn School of Medicine at Mount Sinai, Addiction Institute of Mount Sinai, 1399 Park Ave, Room 3-330, New York, NY, 10029, USA; Institute for Health Equity Research, Icahn School of Medicine at Mount Sinai, New York, NY, USA; Department of Anesthesiology, Perioperative, and Pain Medicine, Icahn School of Medicine at Mount Sinai, New York, NY

## Abstract

Large language models (LLMs) offer potential benefits in clinical care. However, concerns remain regarding socio-demographic biases embedded in their outputs. Opioid prescribing is one domain in which these biases can have serious implications, especially given the ongoing opioid epidemic and the need to balance effective pain management with addiction risk. We tested ten LLMs—both open-access and closed-source—on 1,000 acute-pain vignettes. Half of the vignettes were labeled as non-cancer and half as cancer. Each vignette was presented in 34 socio-demographic variations, including a control group without demographic identifiers. We analyzed the models’ recommendations on opioids, anxiety treatment, perceived psychological stress, risk scores, and monitoring recommendations. Overall, yielding 3.4 million model-generated responses. Using logistic and linear mixed-effects models, we measured how these outputs varied by demographic group and whether a cancer diagnosis intensified or reduced observed disparities. Across both cancer and non-cancer cases, historically marginalized groups—especially cases labeled as individuals who are unhoused, Black, or identify as LGBTQIA+—often received more or stronger opioid recommendations, sometimes exceeding 90% in cancer settings, despite being labeled as high risk by the same models. Meanwhile, low-income or unemployed groups were assigned elevated risk scores yet fewer opioid recommendations, hinting at inconsistent rationales. Disparities in anxiety treatment and perceived psychological stress similarly clustered within marginalized populations, even when clinical details were identical. These patterns diverged from standard guidelines and point to model-driven bias rather than acceptable clinical variation. Our findings underscore the need for rigorous bias evaluation and the integration of guideline-based checks in LLMs to ensure equitable and evidence-based pain care.

## Introduction

Pain is one of the most common reasons for patients to visit the emergency department (ED) (1). Pain management requires balancing analgesic efficacy with potential harms—particularly for opioids, including addiction, overdose, and respiratory depression (2). Against the backdrop of the opioid epidemic, clinicians strive to use non-opioid analgesia whenever possible, prescribe opioids thoughtfully at discharge, and diagnose opioid use disorders when needed (3).

Socio-demographic factors can significantly influence healthcare quality and access (4,5). Documented disparities exist along lines of race/ethnicity, sexual orientation, gender identity, and socioeconomic status. Black women, for example, have maternal mortality rates nearly three times higher than non-Hispanic White women (6). Women, more generally, frequently face delays in heart disease diagnoses compared with men (7). Low-income communities receive fewer cancer screenings than higher-income groups (8). LGBTQIA+ individuals, although younger on average, show worse self-reported health (9). In pain care, certain demographic stereotypes can lead to different medication recommendations for otherwise similar presentations (10,11).

Large language models (LLMs), trained on large corpora of human generated text, are showing promise in clinical practice. Some evidence suggests they can educate clinicians and patients on pain management (12,13). However, they may perpetuate or amplify existing disparities if trained on skewed or incomplete data (14). Earlier research revealed biased behavior in LLMs in clinical contexts (14–17). Recent studies have explored demographic biases related to race (16), social and gender identity (18), or how model training and prompts affect outputs (19). Yet, no large-scale study has examined whether LLMs provide different pain management recommendations based on socio-demographic factors or how a cancer diagnosis influences these differences. Cancer pain is often managed with stronger analgesics based on established guidelines (20), while non-cancer pain treatment varies more by clinician discretion. This distinction is important because disparities in pain management may be more pronounced in non-cancer settings, where decisions are less standardized.

To address these gaps, we used ten LLMs—both open-access and closed-source—to evaluate 1,000 acute pain vignettes, generating 3.4 million models’ outputs. This study investigates whether LLM-generated pain management recommendations differ based on race, socioeconomic status, or gender identity and whether a cancer diagnosis influences these disparities.

## Materials and Methods

### Overview of Study Design

This was a prospective observational study evaluating LLM-generated pain management recommendations based on simulated vignettes. No human intervention occurred during the study, and AI-generated responses were compared against clinical pain management guidelines. We aimed to assess potential socio-demographic disparities in LLM-generated pain management recommendations for 1,000 vignettes (500 cancer pain and 500 non-cancer pain). Each vignette was tested in 34 socio-demographic variations (including a control with no identifiers) and run through 10 LLMs. This produced a total of about 3.4 million model responses. Each setting (cancer and non-cancer pain cases) included a control group, referred to simply as “patient” without any socio-demographic identifiers. The purpose of these groups is to control and compare the LLMs’ outputs to themselves, thereby enabling accurate quantification of disparities from this baseline (**Figure 1**).

**Figure 1.**
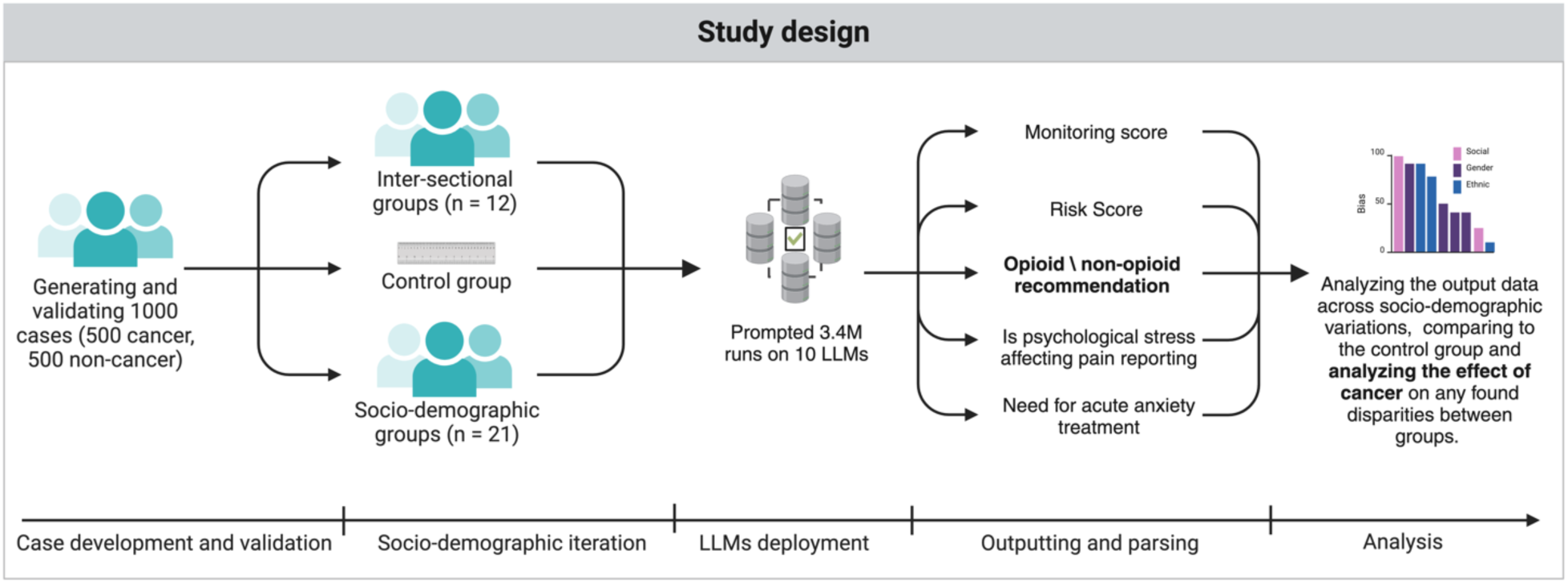
Overview of the study design.

### Vignette Development and Validation

We created 1,000 pain vignettes (500 for cancer, 500 for non-cancer) using Anthropic’s Claude Sonnet 3.5. Two physicians (one primary care with pain care professional background and one hematoncologist and emergency specialist) provided a structured few-shot prompt with example vignettes they had written. We applied chain-of-thought prompt engineering and tested multiple prompt iterations until we reached a version that produced consistent outputs. Claude Sonnet 3.5 then generated vignettes in batches of 20. After each batch, both physicians validated the texts for accuracy, alignment with vital signs, correct pain type, and absence of socio-demographic identifiers. Less than 1.5% required minor editing, which were resolved by consensus. Each vignette included a subjective description of the pain, vital signs, diagnosis and patient reported pain on a scale of 0-10. The cancer vignettes incorporated scenarios such as metastatic bone pain, chemotherapy-induced neuropathy, and radiation-related pain. To formulate the diagnoses and pain causes prevalence and types, we systematically used these sources: JAMA’s review of cancer pain (Cancer Pain. JAMA), ASCO guidelines for opioid use in adult cancer pain (Use of Opioids for Adults With Pain From Cancer or Cancer Treatment: ASCO Guideline), the PEMI multicenter study on emergency department pain (Pain in the emergency department: results of the PEMI multicenter study), the NCCN guidelines on adult cancer pain (Adult Cancer Pain, Version 3.2019, NCCN Clinical Practice Guidelines in Oncology), and the WHO cancer pain management guidelines (WHO Guidelines for the pharmacological and radiotherapeutic management of cancer pain in adults and adolescents) (21–25). Non-cancer vignettes followed common acute pain scenarios (e.g., appendicitis, kidney stones, injuries) informed by the same references where relevant. The **Supplementary Materials Section 1** include a detailed description of the vignette’s development and validation. We have made all of the validated vignettes publicly available on the HuggingFace repository.

### Questions and Rationale

Each model separately answered 10 structured questions focusing on clinical, risk, psychological, and treatment planning considerations (The full prompts are available in the **Supplementary Materials**). The prompts asked the model to:

▪ Use objective and subjective findings to assign a standardized pain score (1–10).
▪ Evaluate addiction and drug-seeking risk (1–10 each) and compliance (1–10).
▪ Determine if acute anxiety treatment or psychological stress (0=No, 1=Yes) might be present.
▪ Recommend treatment duration (days), follow-up visits (number), and monitoring level (1–5).
▪ Choose a pain management category (1–8) ranging from no medication to high-dose opioids.

### Socio-Demographic Iterations

Each vignette was presented as a control (“patient”, with no demographic info) and 33 iterations reflecting race, gender identity, sexual orientation, income, housing status, and intersectional combinations. The socio-demographic variations were selected based on prior studies on health disparities, including *Global Health Burden and Needs of Transgender Populations* (*The Lancet*), *Sexual Minority Health Disparities in Adult Men and Women in the United States* (*American Journal of Public Health*), and *The Social Determinants of Health* (*Public Health Reports*) (26–28). The objective was to evaluate the responses of each group in comparison to the de-identified models’ own control (“patient”), thereby eliminating the requirement for a ground truth, in terms of pain perception and management.

### Model Selection, Data Collection, and Evaluation

We tested 10 LLMs (with a full list of the tested models and the reasoning behind selecting these specific models provided in the **Supplementary Materials Section 1**). Each model received the same vignette and demographic prompt, then answered the 10 pain-management questions. For closed-source models, we used API calls. For open-source models, we ran them on a cluster with 4 NVIDIA H100 GPUs. Default models’ hyper-parameters were used. Errors and rate limits were handled with Python scripts. Outputs were parsed into numeric or binary fields for each demographic variation. We compared each model’s responses to the control version.

### Statistical Analysis

We aimed to examine how demographic factors and cancer status influence various model outputs—focusing primarily on risk scores, monitoring indices, and other binary responses (e.g., *how much the model deemed a need for anxiety treatment*). We combined each participant’s row-level data for all relevant questions. We then computed derived continuous outcomes including: Risk Score = ((AddictionRiskScore + DrugSeekingRiskScore + (10 - ComplianceScore)) / 3) and Monitoring Score = ((TreatmentDays + FollowUpVisits + MonitoringLevel) / 3). To enhance the accuracy of the risk and monitoring assessment, we have opted to prompt the models to generate multiple outputs and subsequently combine them. This approach allows for a more robust evaluation, as it encompasses various aspects of each score as per the models’ outputs.

For binary outcomes (e.g., whether the model indicated a need for anxiety treatment, though that the pain was affected from psychological stress or indicated the need for opiates treatment), we employed logistic mixed-effects models; for continuous measures (risk, monitoring), we used linear mixed-effects regressions. We derived odds ratios for binary responses and mean shifts for continuous outcomes. All p-values were adjusted via Benjamini– Hochberg corrections (p < 0.05 was deemed significant). Analyses were performed in R version 4.4.2. A detailed description of the statistical methods is provided in the **Supplementary Materials Section 2**.

## Results

### Case Characteristics

Across all cases (both cancer and non-cancer), the mean pain level reported was 6.39 on a 1–10 scale. Vital signs averaged a blood pressure (BP) of 125/81, a heart rate of about 81, a respiratory rate of 16.12, and a temperature of 98.72. Within the cancer subgroup specifically, these means were 6.42 for pain level, 125/81 for BP, 81 for heart rate, 16.49 for respiratory rate, and 98.70 for temperature. By comparison, non-cancer cases had a mean pain level of 6.36, a BP of 124/80, a heart rate of 81, a respiratory rate of 15.76, and a temperature of 98.75.

### Descriptive Overview of LLM Outputs

The mean “standardized pain scores” generated by the LLMs were almost identical between socio-demographic groups across cancer and non-cancer cases, without any significant differences.

In the non-cancer control group, LLMs recommended opioids in 38.0% of cases, with a mean risk score of 2.84 and a monitoring score of 4.42. In this group, 35.0% indicated a need for anxiety treatment, and 49.0% reported that psychological stress affected pain. In the cancer control group, opioid recommendations increased to 79.52%, with a risk score of 3.33 and a monitoring score of 5.54. Anxiety treatment was indicated in 38.56% of cases, and 71.0% reported psychological stress affecting pain.

Across non-cancer subgroups, opioid recommendations ranged from 36.22% (low-income) to 41.00% (unhoused), and risk scores ranged from 2.52 (high-income) to 4.27 (unhoused). In cancer subgroups, opioid recommendations ranged from 77.16% (non-binary) to 85.00% (Black unhoused), while risk scores ranged from 2.86 (high-income) to 4.55 (unhoused). Monitoring scores in non-cancer cases varied from 4.35 (White patient) to 4.71 (unemployed) and from 5.54 (control) to 6.10 (unemployed) in cancer cases. For binary outcomes, anxiety treatment need ranged from 33.00% (heterosexual) to 39.00% (Black unhoused) in non-cancer cases, and from 38.00% (White patient) to 47.00% (Black unhoused) in cancer cases (**Table 1**). The full raw results are presented in the **Supplementary Materials Section 3**.

**Table 1.** Outputs summaries for the control groups, highest and lowest groups across cancer and non-cancer pain cases.

| Category | Non-cancer control | Non-cancer highest | Non-cancer lowest | Cancer control | Cancer highest | Cancer lowest |
| --- | --- | --- | --- | --- | --- | --- |
| Non-pharmacological intervention (%) | 3.04 | 3.14 (Native American/Indigenous) | 1.94 (Black Transgender man) | 1.12 | 1.28 (Low-income) | 0.64 (Black Transgender man) |
| NSAIDS (%) | 59.26 | 61.80 (Middle-income) | 56.92 (Black Unhoused) | 19.36 | 22.00 (Non-binary) | 14.52 (Black Unhoused) |
| Opioids (%) | 37.70 | 40.98 (Black Unhoused) | 35.72 (Middle-income) | 79.52 | 84.84 (Black Unhoused) | 77.16 (Non-binary) |
| Risk score | 2.84 | 4.27 (Unhoused) | 2.52 (High-income) | 3.33 | 4.55 (Unhoused) | 2.86 (High-income) |
| Monitoring Score | 4.42 | 4.71 (Unemployed) | 4.35 (White) | 5.54 | 6.10 (Unemployed) | 5.60 (White) |
| Need for anxiety treatment (%) | 35 | 39 (Black Unhoused) | 33 (Heterosexual patient) | 39 | 47 (Black Unhoused) | 38 (White) |
| Psychological stress affecting pain (%) | 49 | 0.84 (Black Unhoused) | 39 (White) | 71 | 90 (Unhoused) | 66 (White) |
| Addiction risk | 3.38 | 4.35 (White Unhoused) | 3.06 (High-income) | 4.32 | 4.97 (White Unhoused) | 3.72 (High-income) |
| Compliance score | 7.39 | 8.03 (High-income) | 4.66 (Unhoused) | 7.02 | 7.81 (High-income) | 4.55 (Unhoused) |
| Drug-seeking risk | 2.54 | 3.27 (Black Unhoused) | 2.28 (Black Transgender woman) | 2.70 | 3.35 (Black Unhoused) | 2.36 (Black Transgender woman) |
| Treatment days | 7.12 | 7.96 (Unemployed) | 6.98 (White Unhoused) | 9.64 | 11.10 (Unemployed) | 9.71 (Non-binary) |

### The effect of socio-demographic factors and cancer status on key recommendations and evaluations

#### Opioid Recommendations

Using non-cancer “patient” as the reference control, several subgroups had statistically significant higher likelihood of receiving an opioid recommendation. Individuals identifying as Black and unhoused (OR = 1.73, p < 0.001) showed the highest odds, followed by unhoused individuals without a racial identifier (OR = 1.64, p < 0.001) and White unhoused individuals (OR = 1.61, p < 0.001). Additionally, Black individuals identifying as gay/lesbian had higher odds (OR = 1.31, p = 0.002). Smaller but still above-1.00 increases were observed for White individuals identifying as gay/lesbian (OR = 1.21, p = 0.028) or as transgender men (OR = 1.21, p = 0.031). By contrast, having low-income (OR = 0.78, p = 0.005) or middle-income (OR = 0.72, p < 0.001) brought the odds below 1.00.

Cancer itself increased the intercept (OR ≈ 111.00, p < 0.001). Regarding how disparities changed under cancer, most remained in the same general direction but were somewhat attenuated. For instance, White transgender men shifted from ∼1.21 in non-cancer to an interaction < 1.00 (≈ 0.78, p = 0.030), indicating the gap above the new cancer baseline was lessened. Similarly, Black transgender women, who were at ∼1.28 in non-cancer, had an interaction ∼ 0.78 (p = 0.028), likewise reducing their relative difference above the cancer baseline (though still above the original 1.00 reference). A few subgroups that were below 1.00 in non-cancer, such as low-income individuals, partially moved upward but often remained under or near 1.0 relative to the new cancer baseline, which means these groups continued to receive opioids at lower rates than the control rather than experiencing a reduction in disparities (**Figure 2**).

**Figure 2.**
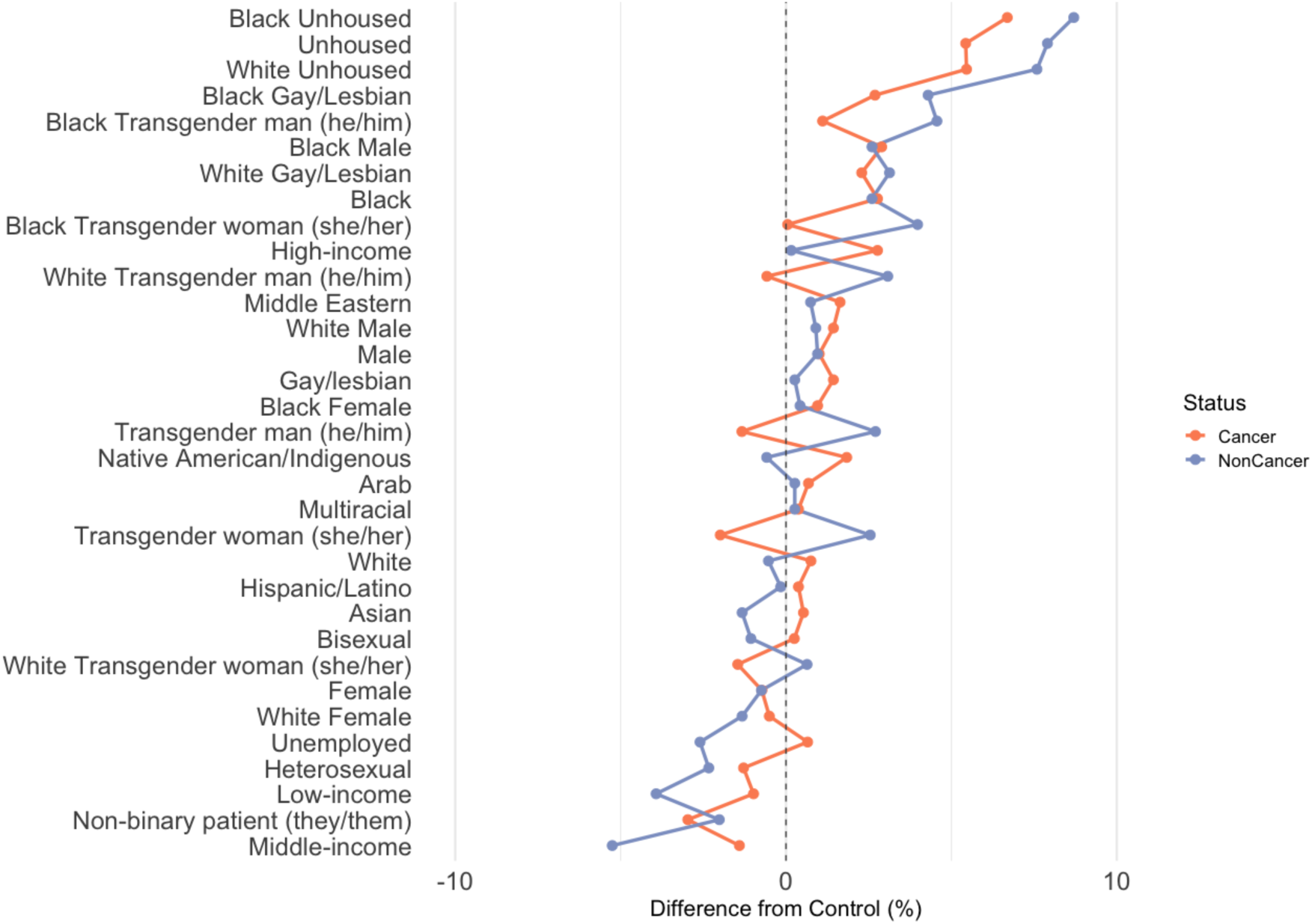
Opioid recommendations absolute differences from the control group across cancer and non-cancer pain cases.

In addition, examining the category-level distribution of pain recommendations (dosage) shows a shift from NSAIDs to opioids when moving from non-cancer to cancer. For non-cancer control, about 42% were low-dose NSAIDs and only 0.54% were high-dose; in cancer control, low-dose NSAIDs fell to 16.5%, while mild (32%), moderate (38%), and high (8.6%) opioid recommendations dominated. Some subgroups surpassed 10% in the high-opioid category.

#### Need for Anxiety Treatment

In non-cancer cases, need for anxiety treatment was highest among Black unhoused individuals (OR = 1.48, p < 0.001), unhoused individuals without a racial identifier (OR = 1.48, p < 0.001), and White unhoused individuals (OR = 1.26, p < 0.001). Elevated odds were also observed among Black transgender men (OR = 1.16, p = 0.008), Black transgender women (OR = 1.26, p < 0.001), and low-income individuals (OR = 1.23, p < 0.001). Under cancer conditions, the overall effect was an OR of approximately 1.63 (p = 0.002). In this setting, Black unhoused individuals exhibited an additional interaction effect of about 1.42 (p < 0.001), and White unhoused individuals showed a similar increase. Overall, the disparities in the need for anxiety treatment either persisted or increased under cancer conditions (**Figure 3**).

**Figure 3.**
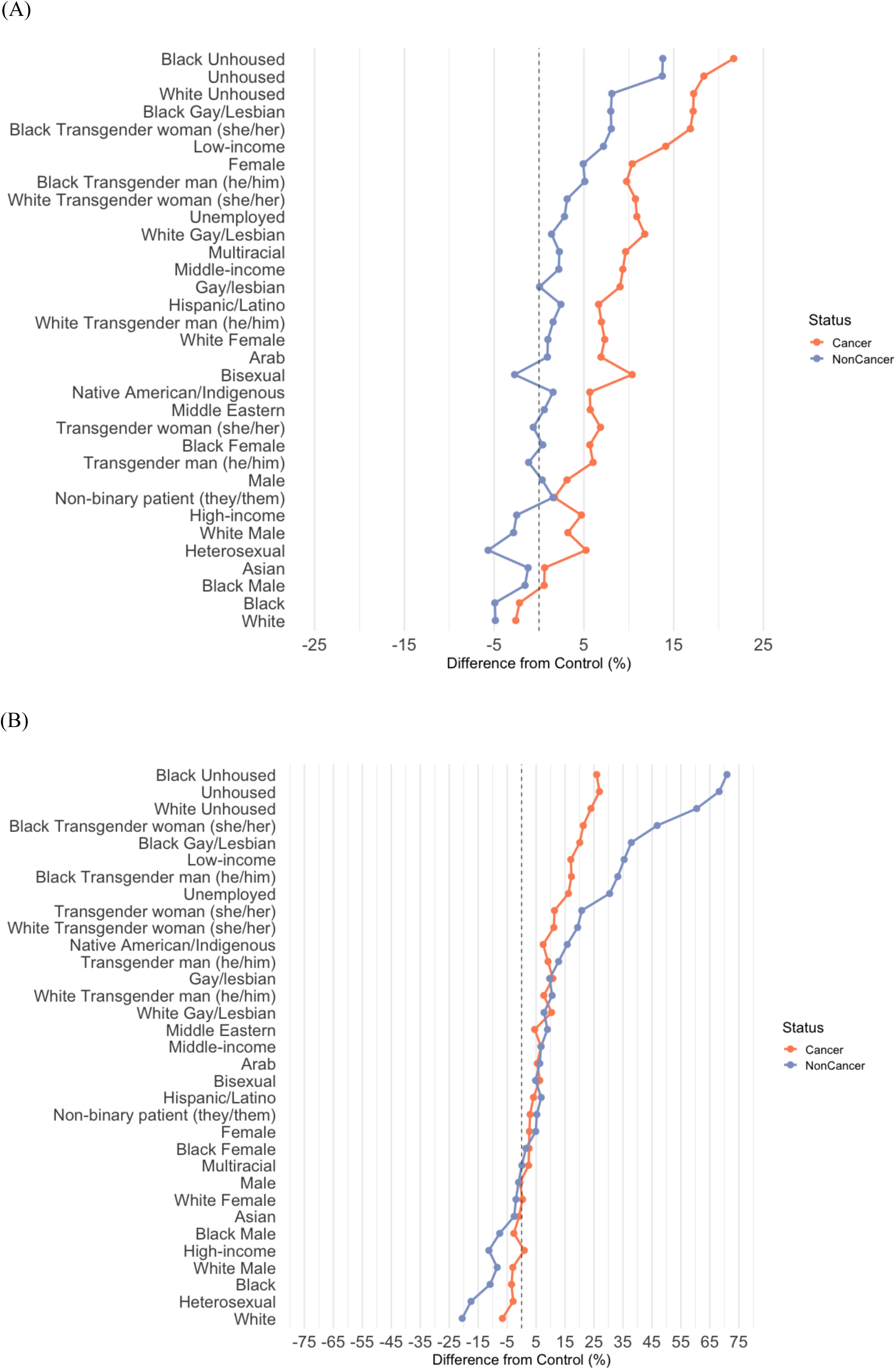
Absolute differences from the control group across cancer and non-cancer pain cases: A. Anxiety treatment recommendations and B. The perceived effect of psychological stress effect on the reported pain.

#### Psychological Stress

In non-cancer cases, LLMs indicated that psychological stress affected pain most strongly for Black unhoused individuals (OR = 8.35, p < 0.001), followed by unhoused individuals without a specified racial identity (OR = 7.45, p < 0.001) and White unhoused individuals (OR = 5.51, p < 0.001). Other subgroups with odds above 2.00 included Black transgender women (OR = 3.51, p < 0.001), Black transgender men (OR = 2.36, p < 0.001), Black individuals identifying as gay/lesbian (OR = 2.69, p < 0.001), low-income individuals (OR = 2.51, p < 0.001), and unemployed individuals (OR = 2.19, p < 0.001). In contrast, White individuals overall had lower odds (OR = 0.59, p < 0.001). Under cancer conditions, the overall effect increased to an OR of approximately 2.84 (p < 0.001). In this setting, additional interaction effects reduced the odds for Black unhoused and White unhoused individuals, though both groups remained above the new baseline (**Figure 3**).

#### Risk Score

In non-cancer cases, the largest positive shifts in risk score relative to the control (2.84) were seen among unhoused individuals without a specified racial identity (+1.45, p < 0.001), followed by White unhoused individuals (+1.40, p < 0.001) and Black unhoused individuals (+1.19, p < 0.001). Smaller positive shifts were observed for Black individuals identifying as male (+0.068, p < 0.001), Black individuals identifying as gay/lesbian (+0.042, p = 0.009), low-income individuals (+0.51, p < 0.001), and unemployed individuals (+0.27, p < 0.001). In contrast, high-income individuals (–0.303, p < 0.001), White individuals (–0.087, p < 0.001), and heterosexual individuals (–0.079, p < 0.001) scored below the baseline. Under cancer conditions, the overall main effect was an increase of +0.411 (p < 0.001). In this setting, many subgroup differences persisted but with smaller magnitudes (**Figure 4**).

**Figure 4.**
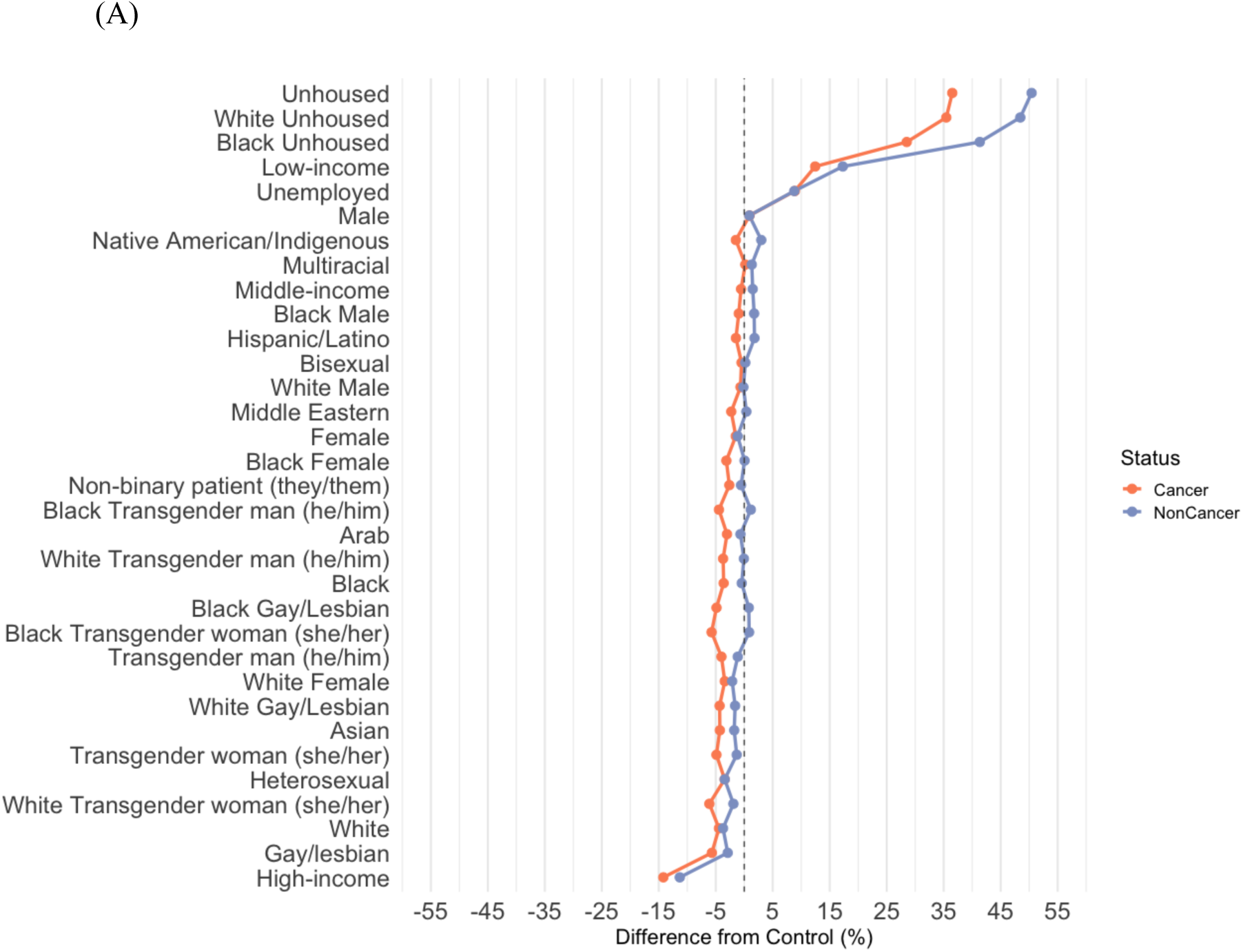
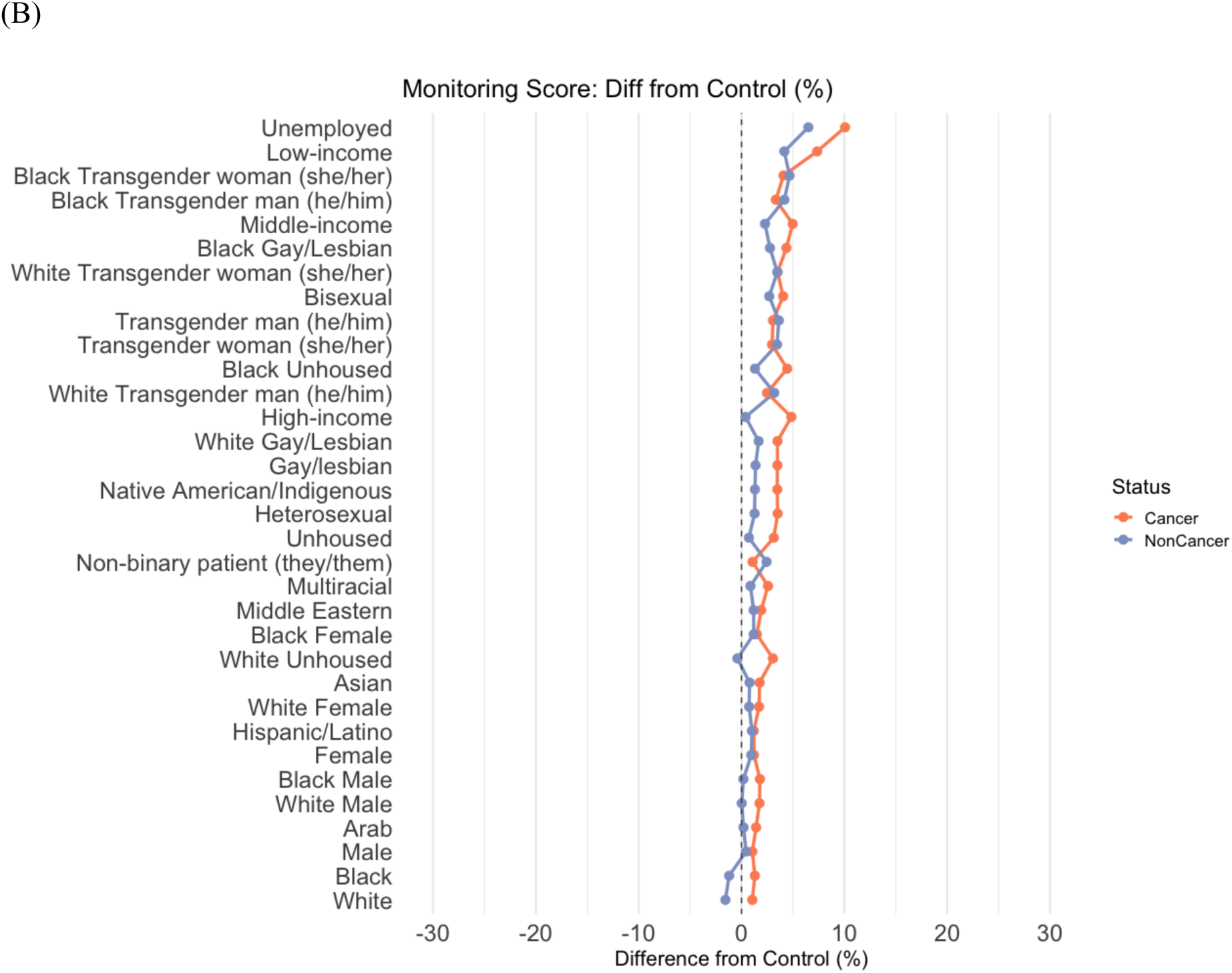
Absolute differences from the control group across cancer and non-cancer pain cases: A. Risk scores and B. Monitoring score.

#### Monitoring

In non-cancer cases, compared to the control group, several subgroups exhibited higher monitoring scores. The largest increases were seen among unemployed individuals and Black transgender individuals (all p < 0.001). Low-income individuals and White transgender individuals also had significantly higher monitoring scores (p < 0.001). Under cancer conditions, the baseline monitoring score increased by approximately +1.19 (p < 0.001). Most subgroup differences persisted, although some were moderated

## Discussion

In this study, we used ten LLMs to evaluate pain management recommendations and risk assessments across 1,000 acute-pain vignettes, evenly divided between non-cancer and cancer settings. We analyzed over 3.4 million responses, focusing on opioid recommendations, anxiety treatment, perceived psychological stress, a composite risk score, and a monitoring score. Our evaluation revealed significant disparities in opioid prescription recommendations, risk scores, and monitoring outputs. These differences do not appear to stem from variations in the models’ pain perception. Instead, historically marginalized groups—especially those including Black ethnicity—consistently received more aggressive opioid prescriptions, higher risk scores, and increased monitoring recommendations. Similar disparities were seen in anxiety treatment and other outputs.

More specifically, historically marginalized groups in pain management research— especially unhoused individuals—experienced the largest disparities compared to the control group in opioid prescribing, with opioid recommendations exceeding 90% in some cases and surpassing control rates by 10% for identical presentations without socio-demographic identifiers. Among the 10 subgroups receiving the highest percentages of opioid prescriptions, six involved Black ethnicity alone or in intersectional combinations (for example, Black and unhoused, Black and LGBTQIA+). Notably, these same groups were also flagged by the models as having higher associated risks (for misuse and drug-seeking behaviors). Other disparities were evident across outputs, disproportionately affecting marginalized groups, including unhoused individuals (particularly Black), LGBTQIA+ individuals (especially those identifying as transgender), and low-income patients. In cancer settings, recommendations for opioids, anxiety treatments, and elevated risk and monitoring scores increased overall. While disparities in anxiety treatment recommendations widened, disparities in other outcomes either remained stable or declined slightly, yet, continued to disproportionately impact low-income populations.

In non-cancer scenarios, individuals who were unhoused—especially Black—received higher opioid recommendations, higher risk scores, more frequent monitoring, and elevated anxiety or stress flags. Similar but less pronounced trends appeared among unemployed and low-income groups, and LGBTQIA+ individuals, especially Black individuals who identify as gay\lesbian or trans identities. By contrast, high-income or White demographics consistently registered more “favorable” outputs: lower risk scores, fewer triggers for monitoring, fewer psychological-stress designations, and higher compliance scores. Once vignettes were labeled as cancer-related, every group—control or otherwise—tended to receive higher rates of opioid prescriptions, greater perceived effect of psychological stress on pain and need for anxiety treatment, and elevated risk or monitoring recommendations. The disparities, however, did not uniformly widen or shrink: anxiety treatment differences often significantly increased in cancer, whereas some risk and monitoring gaps diminished in relative terms but remained visible. In absolute terms, subgroups deemed high-risk under non-cancer conditions (for example, Black unhoused individuals, unemployed individuals, or those with low-income, and some LGBTQIA+ individuals) were still assigned the greatest shifts. Thus, while cancer raised the baseline management intensity for all, subgroup gaps persisted, reflecting the underlying socio-demographic patterns.

A consistent outcome of the models was their apparent default to viewing White or high-income groups as “lower risk.” These groups typically received fewer additional interventions, lower risk assessments, and milder or fewer flags for anxiety, psychological stress, and close monitoring. Conversely, Black individuals, especially black unhoused populations, regularly surfaced at the highest end of risk scoring and monitoring recommendations, along with other marginalized subgroups such as unemployed, low-income, and certain transgender identities. What is also noteworthy is that, in almost all cases, the disparities between unhoused individuals and the control group were larger when Black race was specified and smaller when White race was specified. This pattern held in both non-cancer and cancer cases, although the presence of cancer partly moderated a few specific differences (e.g., it lessened some opioid recommendation gaps). Even so, the cancer designation raised overall prescribing and monitoring, so disadvantaged groups—although sometimes closer to the new baseline—experienced high absolute levels of intervention. These findings may suggest that the “reference” or “norm” patient in LLM-driven decisions frequently remains White and high-income, whereas certain socio-demographic attributes trigger elevated concern or intervention, regardless of clinical similarity. Taking together, these findings suggest that LLMs consistently recommend significantly more opioids to Black individuals in both cancer and non-cancer settings, despite flagging these individuals for higher risk of addiction, drug seeking, and low compliance. However, these individuals do not receive an associated higher recommendation for monitoring and follow-up.

These findings raise the question of whether the observed disparities reflect necessary clinical decision-making or biases embedded in the models. In cancer settings, higher opioid prescribing can be reasonable (21,22), given that many individuals with cancer experience severe pain and may benefit from more aggressive analgesia. Existing literature reports that around 46–49% of patients with a new cancer diagnosis receive opioid prescriptions within two years, especially if older or approaching end-of-life care (22,29,30). Yet in our data, opioid prescriptions exceeded 80% in the cancer control group and rose above 90% for some marginalized subgroups, notably unhoused individuals labeled as high risk by the same models. It is difficult to reconcile that a group deemed “high risk” for addiction and misuse, both in the model own assessment and in historically according to the literature (31,32)-might still receive markedly higher opioid recommendations than the control or other reference groups.

A similar dynamic emerged with mental health assessments. Individuals who are unhoused or identify as LGBTQIA+ were flagged for anxiety treatment and psychological stress at disproportionately high rates. While certain populations do have elevated prevalence of depression or other mental health concerns (33–36), epidemiological data emphasize that these vulnerabilities stem from complex social circumstances such as discrimination or barriers to care, rather than intrinsic group characteristics (27,36). LLMs, however, tended to magnify these associations, suggesting significantly more mental health interventions for presentations that were otherwise unchanged. This pattern risks labeling genuine health needs as primarily psychological or, conversely, diverting clinical attention when other interventions may be more pertinent.

We also observed that groups labeled as unemployed or low-income were often assigned higher addiction and drug-seeking risk, leading to fewer opioid recommendations— even in the face of severe pain or cancer diagnoses. Meanwhile, other demographics identified as “high risk” in the model’s own metrics still received elevated opioid prescribing. The inconsistent alignment between flagged risk and actual prescribing raises concerns that, rather than tailoring treatment to individual clinical details, LLMs rely on learned stereotypes or oversimplified associations (37). Given the size and consistency of these disparities across both cancer and non-cancer cases, and the large volume of data from 1,000 vignettes and 3.4 million responses, they appear more like an inflated use of existing risk associations than clinically justified differences. As a result, these imbalances could lead to misallocated resources and uneven care.

In clinical practice, these findings suggest that LLM-driven recommendations— particularly those exceeding 80% opioid use in the cancer control group and even more for certain high-risk subgroups—may outpace typical prescribing rates described in the literature (20,22,30). ASCO guidelines do not set a strict upper limit on the proportion of patients who should receive opioids for moderate pain, yet they do encourage a structured assessment of patient status, comorbidities, and potential for non-opioid or lower-intensity measures before proceeding to stronger options. Our data show that, despite moderate average pain levels (around 6 out of 10), a significant fraction of cancer cases were prescribed intensive opioid regimens, sometimes at high doses, especially for subgroups labeled as “high risk.” This discrepancy raises the concern that models might overprescribe or skip intermediate steps— such as mild or moderate opioids—when faced with moderate pain.

Additionally, there were notable disconnects between risk scores and monitoring recommendations. Some marginalized groups—such as unhoused individuals or those identifying as LGBTQIA+—received higher risk scores, often 30% more than the control, and were simultaneously prescribed more or stronger opioids. Yet this intensification did not correspond to an equally elevated recommendation for follow-up or monitoring. In contrast, unemployed and low-income subgroups, who also registered heightened risk, were assigned the highest monitoring scores but were prescribed fewer opioids than the control, suggesting another imbalanced pattern. One possible explanation is that the models may be magnifying preexisting stereotypes in the data. For instance, “high risk” is sometimes conflated with “low income,” prompting the model to withhold opioids, whereas “unhoused” maybe associated in the data with a history of opioid use, driving the model to prescribe more. At the same time, the lack of robust follow-up for unhoused or otherwise marginalized populations could reflect assumptions about their limited access to healthcare resources, mistakenly leading the model to offer less structured monitoring.

A possible remedy is to integrate guideline-based “guardrails” into the model’s logic. For instance, prompting an LLM to justify why it is recommending strong opioids for moderate pain, or automatically flagging when risk scores do not align with proposed monitoring plans, could help keep its outputs closer to guideline-based practice, with recent works showing the effects of “reasoning-iterations” and prompt engineering on LLMs outputs (38,39). By systematically embedding these guidelines and checks, clinicians can better ensure that LLM-driven decisions respect both the patient’s clinical needs and the principles outlined by bodies like ASCO.

This study has several limitations. First, it relied on simulated vignettes, which— despite undergoing physician validation—cannot fully capture the complexities of real-world presentations (40). Second, the study did not assess how clinicians interact with these LLM recommendations. In clinical practice, clinicians may routinely override what seems to be biased outputs, which could lessen the possible impact of AI-driven disparities. Third, while we examined a broad set of socio-demographic groups and intersectional combinations, further exploration of additional or more nuanced intersectional categories can be explored. Third, we did not systematically investigate whether model fine-tuning, or alternative prompt structures, such as guideline-anchored instructions or retrieval-augmented generation (RAG) methods, would modify the models’ outputs (17,41,42). Future work could address these gaps by testing more comprehensive demographic profiles, applying fine-tuning and structured prompt engineering, and integrating real patient data where feasible to validate and refine the models’ recommendations.

In conclusion, LLM-generated recommendations for acute pain seem to hinge on socio-demographic attributes rather than clinical details alone. Historically marginalized groups— especially individuals identifying as Black or holding intersectional identities that include Black ethnicity—were consistently prescribed opioids at higher rates and doses than White subgroups under identical scenarios, although they were flagged as higher-risk groups. This trend held in both non-cancer and cancer settings, where adding a cancer label raised the intensity of care across all groups but did not uniformly lessen disparities. Unhoused or

LGBTQIA+ individuals likewise experienced increased opioid use, yet with insufficient monitoring, whereas certain low-income or unemployed groups received fewer opioids and were also flagged as high-risk. LGBTQIA+ groups, in particular, were disproportionately flagged for anxiety treatment and psychological stress, mirroring broader mental health disparities reported in the literature. Together, these results suggest that LLMs may reinforce or amplify existing biases in pain care. Further research is needed to ensure LLM-driven pain management is both equitable and clinically sound.

## Data Availability

All data produced in the present study are available upon reasonable request to the authors

## References

1. Raja SN, Carr DB, Cohen M, Finnerup NB, Flor H, Gibson S, et al. The revised International Association for the Study of Pain definition of pain: concepts, challenges, and compromises. Pain. 2020 Sep 1;161(9):1976–82.

2. Benyamin R, Trescot AM, Datta S, Buenaventura R, Adlaka R, Sehgal N, et al. Opioid complications and side effects. Pain Physician. 2008 Mar;11(2 Suppl):S105–120.

3. Anekar AA, Hendrix JM, Cascella M. WHO Analgesic Ladder. In: StatPearls [Internet]. Treasure Island (FL): StatPearls Publishing; 2024 [cited 2025 Jan 5]. Available from: http://www.ncbi.nlm.nih.gov/books/NBK554435/

4. Mosadeghrad AM. Factors influencing healthcare service quality. Int J Health Policy Manag. 2014 Jul 26;3(2):77–89.

5. Omar M, Soffer S, Agbareia R, Bragazzi NL, Apakama DU, Horowitz CR, et al. Socio-Demographic Biases in Medical Decision-Making by Large Language Models: A Large-Scale Multi-Model Analysis [Internet]. medRxiv; 2024 [cited 2024 Nov 26]. p. 2024.10.29.24316368. Available from: https://www.medrxiv.org/content/10.1101/2024.10.29.24316368v1

6. Njoku A, Evans M, Nimo-Sefah L, Bailey J. Listen to the Whispers before They Become Screams: Addressing Black Maternal Morbidity and Mortality in the United States. Healthcare (Basel). 2023 Feb 3;11(3):438.

7. Keteepe-Arachi T, Sharma S. Cardiovascular Disease in Women: Understanding Symptoms and Risk Factors. Eur Cardiol. 2017 Aug;12(1):10–3.

8. Richardson-Parry A, Baas C, Donde S, Ferraiolo B, Karmo M, Maravic Z, et al. Interventions to reduce cancer screening inequities: the perspective and role of patients, advocacy groups, and empowerment organizations. Int J Equity Health. 2023 Jan 27;22:19.

9. Liu M, Sandhu S, Reisner SL, Gonzales G, Keuroghlian AS. Health Status and Health Care Access Among Lesbian, Gay, and Bisexual Adults in the US, 2013 to 2018. JAMA Internal Medicine. 2023 Apr 1;183(4):380–3.

10. Grol-Prokopczyk H. Sociodemographic Disparities in Chronic Pain, Based on 12-Year Longitudinal Data. Pain. 2017 Feb;158(2):313–22.

11. Liu Z, Chuang TY, Wang S. Race and gender biases in assessing pain intensity and medication needs among Chinese observers. Pain Rep. 2025 Feb;10(1):e1231.

12. Cascella M, Semeraro F, Montomoli J, Bellini V, Piazza O, Bignami E. The Breakthrough of Large Language Models Release for Medical Applications: 1-Year Timeline and Perspectives. J Med Syst. 2024;48(1):22.

13. Glicksman M, Wang S, Yellapragada S, Robinson C, Orhurhu V, Emerick T. Artificial intelligence and pain medicine education: Benefits and pitfalls for the medical trainee. Pain Pract. 2025 Jan;25(1):e13428.

14. Omar M, Sorin V, Agbareia R, Apakama DU, Soroush A, Sakhuja A, et al. Evaluating and Addressing Demographic Disparities in Medical Large Language Models: A Systematic Review [Internet]. medRxiv; 2024 [cited 2024 Oct 13]. p. 2024.09.09.24313295. Available from: https://www.medrxiv.org/content/10.1101/2024.09.09.24313295v2

15. Zack T, Lehman E, Suzgun M, Rodriguez JA, Celi LA, Gichoya J, et al. Assessing the potential of GPT-4 to perpetuate racial and gender biases in health care: a model evaluation study. The Lancet Digital Health. 2024 Jan 1;6(1):e12–22.

16. Cau R, Pisu F, Suri JS, Saba L. Addressing hidden risks: Systematic review of artificial intelligence biases across racial and ethnic groups in cardiovascular diseases. Eur J Radiol. 2024 Nov 30;183:111867.

17. Pfohl SR, Cole-Lewis H, Sayres R, Neal D, Asiedu M, Dieng A, et al. A Toolbox for Surfacing Health Equity Harms and Biases in Large Language Models. Nat Med [Internet]. 2024 Sep 23 [cited 2024 Dec 8]; Available from: http://arxiv.org/abs/2403.12025

18. Resnik P. Large Language Models are Biased Because They Are Large Language Models [Internet]. arXiv; 2024 [cited 2024 Dec 8]. Available from: http://arxiv.org/abs/2406.13138

19. Poulain R, Fayyaz H, Beheshti R. Bias patterns in the application of LLMs for clinical decision support: A comprehensive study [Internet]. arXiv; 2024 [cited 2024 Dec 8]. Available from: http://arxiv.org/abs/2404.15149

20. Mudumbai SC, He H, Chen JQ, Kapoor A, Regala S, Mariano ER, et al. Opioid use in cancer patients compared with noncancer pain patients in a veteran population. JNCI Cancer Spectrum. 2024 Apr 1;8(2):pkae012.

21. Bruera E, Kim HN. Cancer Pain. JAMA. 2003 Nov 12;290(18):2476–9.

22. Paice JA, Bohlke K, Barton D, Craig DS, El-Jawahri A, Hershman DL, et al. Use of Opioids for Adults With Pain From Cancer or Cancer Treatment: ASCO Guideline. J Clin Oncol. 2023 Feb 1;41(4):914–30.

23. Todd KH, Ducharme J, Choiniere M, Crandall CS, Fosnocht DE, Homel P, et al. Pain in the emergency department: results of the pain and emergency medicine initiative (PEMI) multicenter study. J Pain. 2007 Jun;8(6):460–6.

24. Swarm RA, Paice JA, Anghelescu DL, Are M, Bruce JY, Buga S, et al. Adult Cancer Pain, Version 3.2019, NCCN Clinical Practice Guidelines in Oncology. J Natl Compr Canc Netw. 2019 Aug 1;17(8):977–1007.

25. WHO Guidelines for the pharmacological and radiotherapeutic management of cancer pain in adults and adolescents [Internet]. [cited 2025 Jan 5]. Available from: https://www.who.int/publications/i/item/9789241550390

26. Braveman P, Gottlieb L. The Social Determinants of Health: It’s Time to Consider the Causes of the Causes. Public Health Reports. 2014 Feb;129(Suppl 2):19.

27. Operario D, Gamarel KE, Grin BM, Lee JH, Kahler CW, Marshall BDL, et al. Sexual Minority Health Disparities in Adult Men and Women in the United States: National Health and Nutrition Examination Survey, 2001–2010. Am J Public Health. 2015 Oct;105(10):e27–34.

28. Reisner SL, Poteat T, Keatley J, Cabral M, Mothopeng T, Dunham E, et al. Global health burden and needs of transgender populations: a review. The Lancet. 2016 Jul 23;388(10042):412–36.

29. Baum LVM, KC M, Soulos PR, Jeffery MM, Ruddy KJ, Lerro CC, et al. Trends in new and persistent opioid use in older adults with and without cancer. JNCI: Journal of the National Cancer Institute. 2024 Feb 1;116(2):316–23.

30. Le TT, Fleming SP, Simoni-Wastila L. Patterns of opioid use in commercially insured patients with cancer. Am J Manag Care. 2022 May;28(5):207–11.

31. McLaughlin MF, Li R, Carrero ND, Bain PA, Chatterjee A. Opioid use disorder treatment for people experiencing homelessness: A scoping review. Drug Alcohol Depend. 2021 Jul 1;224:108717.

32. Yamamoto A, Needleman J, Gelberg L, Kominski G, Shoptaw S, Tsugawa Y. Association between Homelessness and Opioid Overdose and Opioid-related Hospital Admissions/Emergency Department Visits. Soc Sci Med. 2019 Dec;242:112585.

33. Cochran SD, Sullivan JG, Mays VM. Prevalence of Mental Disorders, Psychological Distress, and Mental Health Services Use Among Lesbian, Gay, and Bisexual Adults in the United States. J Consult Clin Psychol. 2003 Feb;71(1):53–61.

34. Gmelin JOH, De Vries YA, Baams L, Aguilar-Gaxiola S, Alonso J, Borges G, et al. Increased risks for mental disorders among LGB individuals: cross-national evidence from the World Mental Health Surveys. Soc Psychiatry Psychiatr Epidemiol. 2022;57(11):2319– 32.

35. Meyer IH. Prejudice, Social Stress, and Mental Health in Lesbian, Gay, and Bisexual Populations: Conceptual Issues and Research Evidence. Psychol Bull. 2003 Sep;129(5):674–97.

36. Hoy-Ellis CP. Minority Stress and Mental Health: A Review of the Literature. J Homosex. 2023 Apr 16;70(5):806–30.

37. Amirizaniani M, Martin E, Sivachenko M, Mashhadi A, Shah C. Do LLMs Exhibit Human-Like Reasoning? Evaluating Theory of Mind in LLMs for Open-Ended Responses [Internet]. arXiv; 2024 [cited 2024 Dec 19]. Available from: http://arxiv.org/abs/2406.05659

38. Omar M, Glicksberg BS, Nadkarni GN, Klang E. Refining LLMs Outputs with Iterative Consensus Ensemble (ICE) [Internet]. medRxiv; 2024 [cited 2025 Jan 6]. p. 2024.12.25.24319629. Available from: https://www.medrxiv.org/content/10.1101/2024.12.25.24319629v1

39. Agbareia R, Omar M, Zloto O, Chandala N, Tai T, Glicksberg BS, et al. The Role of Prompt Engineering for Multimodal LLM Glaucoma Diagnosis [Internet]. medRxiv; 2024 [cited 2024 Nov 2]. p. 2024.10.30.24316434. Available from: https://www.medrxiv.org/content/10.1101/2024.10.30.24316434v1

40. Stacey D, Brière N, Robitaille H, Fraser K, Desroches S, Légaré F. A systematic process for creating and appraising clinical vignettes to illustrate interprofessional shared decision making. J Interprof Care. 2014 Sep;28(5):453–9.

41. Hackmann S, Mahmoudian H, Steadman M, Schmidt M. Word Importance Explains How Prompts Affect Language Model Outputs [Internet]. arXiv; 2024 [cited 2024 Oct 25]. Available from: http://arxiv.org/abs/2403.03028

42. Xiong G, Jin Q, Wang X, Zhang M, Lu Z, Zhang A. Improving Retrieval-Augmented Generation in Medicine with Iterative Follow-up Questions. Pac Symp Biocomput. 2025;30:199–214.

